# Overview of Direct Oral Anticoagulation Trends in the Bronx: Patient and Systemic Factors Contributing to Medication Nonadherence

**DOI:** 10.1101/2025.04.28.25326611

**Authors:** Sujoy Khasnavis, Jee-Young Moon, Daniel Lerner, Ephraim Leiderman

## Abstract

**Introduction:** Anticoagulation nonadherence has historically been attributed to myriad factors in patient populations across the world. While direct oral anticoagulants (DOACs) have demonstrated advantages such as fewer interactions with medications and freedom from routine lab monitoring compared to other anticoagulants, nonadherence persists and the underlying contributors vary by geography.

**Methods:** This retrospective review used all available electronic medical records on patients receiving active DOAC therapy from primary care centers in the Bronx between 2017-2024. Adherence and nonadherence groups were determined by prescription fill status and provider documentation. The two groups were compared by age, gender, race, ethnicity, insurance type, diagnosis, pharmacy type, employment status, comorbidities, home medications, and primary language. Univariable and multivariable logistic regressions were applied between the groups and categories. P values < 0.05 were deemed significant.

**Results:** The cohort had 938 patients with nonadherence reported in 227 (24.2%) patients. Nonadherence was more common in younger patients (OR 1.20, CI 1.04-1.39, p=0.010) and in males (OR 1.44, CI 1.04-1.99, p=0.026). It was more common in black Non Hispanics (OR 1.98, CI 1.17-3.47, p=0.014) and in the employed (OR 1.92, CI 1.09-3.57, p=0.032). It was also more frequent among patients prescribing from hospital pharmacies (OR 2.63, CI 1.89-3.57, p=0.001) and in English speakers (OR 1.69, CI 1.11-2.63, p=0.016).

**Conclusion:** DOAC nonadherence in the Bronx is significantly associated with younger age, male gender, black Non Hispanic origins, employment, hospital pharmacy prescriptions, and English primary language. Other categories were borderline significant or not significant and warrant future investigation.

## Introduction

Anticoagulation (AC) nonadherence has historically been attributed to myriad factors in patient populations across the world. Among the different classes of ACs, warfarin and heparin have been shown to have their own sets of benefits including superiority in mechanical valves and valvular atrial fibrillation for the former and easy reversibility prior to surgery for the latter. In recent years, the advent of direct oral ACs (DOACs) has provided several key advantages over vitamin K and antithrombin antagonists. Among the advantages is the absence of troublesome interactions with medications, dietary components, and supplements. Additionally, DOACs do not require routine laboratory monitoring to ensure that coagulation levels are consistently therapeutic. Lastly, DOACs do not pose a risk for thrombosis, an issue often seen with warfarin at subtherapeutic international normalized ratios (INRs) and seldom with heparin products in cases of heparin induced thrombocytopenia (HIT). Despite these advances, however, nonadherence to AC therapy persists around the world and while the clinical obstacles in AC therapy have been offset with DOACs, demographic level obstacles continue to diminish the effective utilization of DOACs. As underlying contributors vary by geography, the goal of this retrospective review is to highlight the patient and systemic characteristics associated with DOAC nonadherence in the North Bronx.

## Methods

### Data acquisition

This review utilized electronic medical records from hospital centers in the Bronx. Records were obtained from Epic database. Patients receiving outpatient primary care services and DOAC therapy as of 2024 were included in the analysis. Adherence and nonadherence were determined based on prescription fill status and healthcare provider documentation of medication intake patterns. Nonadherence was defined as pharmacy documentation of medication filling status as “ not filled” or “ first fill/refill not completed” on the most recent DOAC prescription. Nonadherence was also defined as provider documentation of the terms “ not adherent”, “ nonadherent”, “ not compliant”, or “ noncompliant” .

Data were collected from Epic database on age, gender, race, ethnicity, insurance type, indication for DOAC, pharmacy type, employment status, number of comorbidities, number of home medications, primary language, and mental health issues. Indications for DOAC included deep vein thrombosis (DVT), pulmonary embolism (PE), atrial fibrillation/flutter (AF), and other hypercoagulable conditions.

### Statistical analysis

Demographic and clinical variables were summarized by mean standard deviation (SD) for continuous variables and count (%) for categorical variables according to adherence status; they were compared using t-test or Chi-squared test. The odds ratios (OR) with confidence interval (CI) for non-adherence were calculated using a univariable logistic regression. A multivariable logistic regression was fitted for simultaneous adjustment of variables. A backward model selection with Akaike Information Criterion (AIC) was applied to the most significant associations. P values < 0.05 were considered significant and between 0.05 to 0.1 were considered borderline significant.

## Results

Among 938 patients on active DOAC therapy in the Bronx, 227 (24.2%) were identified as nonadherent. Nonadherence was reported by pharmacy in 69 (30.4%) patients and by providers in the remaining 158 (69.6%) patients. Patients ranged from the ages of 25 to 97. Males comprised 434 (46.3%) of the total cohort. Diagnosis types included atrial fibrillation, atrial flutter, deep vein thrombosis, pulmonary embolism, and other hypercoagulable conditions requiring AC.

Tables 1 and 2 show the univariate association with nonadherence. Most variables showed significant associations except for diagnosis type and number of comorbidities. Mental health disorders were reported in less than 1% of both groups and therefore not included in the analysis. Overall, nonadherence was more common among young, male, and employed patients. Compared to non-Hispanic Whites, non-Hispanic Blacks or Hispanics exhibited lower medication adherence. In contrast, Spanish-speaking individuals were more adherent than English speakers. These contrasting associations between race/ethnicity and language with nonadherence will be examined in depth later. The pharmacy type showed the strongest association with nonadherence - the patients who receive prescriptions at hospital pharmacy had three times the odds of being nonadherent compared to those who receive from community/retail pharmacy.

**Table 1.** Comparison of demographic and clinical variables between nonadherence and adherence.

| Category |  | Nonadherent (N=227) | Adherent (N=711) | P-value |
| --- | --- | --- | --- | --- |
| Age |  | 66.3 (12.1) | 70.5 (11.8) | < 0.001 |
| Gender | Female | 103 (45.4) | 401 (56.4) | <0.005 |
|  | Male | 124 (54.6) | 310 (43.6) |  |
| Race ethnicity |  |  |  | <0.001 |
| White/Not Hispanic or Latinx |  | 21 (9.3) | 106 (14.9) |  |
| Black/Not Hispanic or Latinx |  | 122 (54) | 265 (37.3) |  |
| Other/Not Hispanic or Latinx |  | 18 (8) | 100 (14.1) |  |
| White/Hispanic or Latinx |  | 19 (8.4) | 47 (6.6) |  |
| Black/Hispanic or Latinx |  | 10 (4.4) | 35 (4.9) |  |
| Other/Hispanic or Latinx |  | 36 (15.9) | 158 (22.2) |  |
| Language | English | 179 (78.9) | 468 (65.8) | <0.001 |
|  | Spanish | 33 (14.5) | 159 (22.4) |  |
|  | Other | 15 (6.6) | 84 (11.8) |  |
| Employment | Employed | 198 (87.2) | 557 (78.3) | <0.001 |
|  | Retired | 15 (6.6) | 124 (17.4) |  |
|  | Unknown | 14 (6.2) | 30 (4.2) |  |
| Insurance type | Private | 25 (11) | 56 (7.9) | <0.01 |
|  | Medicaid | 155 (68.3) | 441 (62) |  |
|  | Medicare | 47 (20.7) | 214 (30.1) |  |
| Pharmacy type | Hospital | 108 (47.6) | 165 (23.2) | <0.001 |
|  | Community/retail | 119 (52.4) | 546 (76.8) |  |
| Diagnosis | Atrial fibrillation/flutter | 141 (62.1) | 491 (69.1) | >0.116 |
|  | Deep vein thrombosis | 64 (28.2) | 146 (20.5) |  |
|  | Pulmonary embolism | 16 (7) | 56 (7.9) |  |
|  | Other | 6 (2.6) | 18 (2.5) |  |
| Number of comorbidities | <5 | 46 (20.3) | 128 (18) | >0.294 |
|  | 5+ | 100 (44.1) | 288 (40.5) |  |
|  | 10+ | 81 (35.7) | 295 (41.5) |  |
| Number of medications | <5 | 43 (18.9) | 88 (12.4) | <0.025 |
|  | 5+ | 96 (42.3) | 296 (41.6) |  |
|  | 10+ | 88 (38.8) | 327 (46) |  |

Values are mean (SD) or count (%). P-value by t-test or Chi-squared test.

**Table 2.** Unadjusted logistic regression of demographic and clinical variables with nonadherence using OR and CI.

| Category | Nonadherence OR | 95% CI | P-value | P-value (overall) |
| --- | --- | --- | --- | --- |
| Age (per 10 years) | 0.75 | 0.66-0.85 | 0.001 | 0.001 |
| Gender |  |  |  |  |
| Female (ref) | 1.56 | 1.15-2.11 | 0.004 | 0.004 |
| Male |  |  |  |  |
| Race ethnicity |  |  |  |  |
| White/Not Hispanic Latinx (ref) |  |  |  |  |
| Black/Not Hispanic or Latinx | 2.32 | 1.41-3.98 | 0.001 | 0.001 |
| Other/Not Hispanic or Latinx | 0.91 | 0.45-1.8 | 0.784 |  |
| White/Hispanic or Latinx | 2.04 | 1-4.16 | 0.049 |  |
| Black/Hispanic or Latinx | 1.44 | 0.6-3.3 | 0.395 |  |
| Other/Hispanic or Latinx | 1.15 | 0.64-2.11 | 0.643 |  |
| Language |  |  |  |  |
| English (ref) |  |  |  |  |
| Spanish | 0.54 | 0.35-0.81 | 0.004 | 0.001 |
| Other | 0.47 | 0.25-0.81 | 0.010 |  |
| Employment |  |  |  |  |
| Employed (ref) |  |  |  |  |
| Retired | 0.34 | 0.19-0.58 | 0.001 | 0.001 |
| Unknown | 1.31 | 0.66-2.48 | 0.415 |  |
| Insurance_type |  |  |  |  |
| Private (ref) |  |  |  |  |
| Medicaid | 0.79 | 0.48-1.32 | 0.354 | 0.013 |
| Medicare | 0.49 | 0.28-0.88 | 0.014 |  |
| Pharmacy_type |  |  |  |  |
| Hospital (ref) | 0.33 | 0.24-0.46 | 0.001 | 0.001 |
| Community/retail |  |  |  |  |
| Diagnosis |  |  |  |  |
| Atrial fibrillation/flutter (ref) |  |  |  |  |
| Deep vein thrombosis | 1.53 | 1.07-2.16 | 0.017 | 0.127 |
| Pulmonary embolism | 0.99 | 0.54-1.75 | 0.986 |  |
| Other | 1.16 | 0.41-2.83 | 0.757 |  |
| Number_of_comorbidities |  |  |  |  |
| <5 (ref) |  |  |  |  |
| 5+ | 0.97 | 0.65-1.46 | 0.868 | 0.291 |
| 10+ | 0.76 | 0.51-1.16 | 0.206 |  |
| Number_of_medications |  |  |  |  |
| <5 (ref) |  |  |  |  |
| 5+ | 0.66 | 0.43-1.03 | 0.062 | 0.029 |
| 10+ | 0.55 | 0.36-0.85 | 0.007 |  |

In the multivariable logistic regression (Table 3), independent variables associated with nonadherence were male vs female gender (OR 1.42, CI 1.02-1.98, p = 0.037), employment vs retirement (OR 1.89, CI 1.06-3.57, p = 0.037), and hospital vs retail pharmacy prescriptions (OR 2.7, CI 1.92-3.85, p = 0.001). Age, language, and race/ethnicity showed borderline significance (0.05 < p < 0.1). Insurance type, number of comorbidities, and number of home medications did not show significant associations after adjusting for other variables.

**Table 3.** Multivariable logistic regression of demographic and clinical variables with nonadherence using OR and CI.

| Category | Nonadherence OR | 95% CI | P-value | P-value (overall) |
| --- | --- | --- | --- | --- |
| Age (per 10 years) | 0.87 | 0.74-1.01 | 0.072 | 0.072 |
| Gender |  |  |  |  |
| Female (ref) | 1.42 | 1.02-1.98 | 0.037 | 0.037 |
| Male |  |  |  |  |
| Race_ethnicity |  |  |  |  |
| White/Not Hispanic Latinx (ref) |  |  |  |  |
| Black/Not Hispanic or Latinx | 1.75 | 0.97-3.26 | 0.069 | 0.037 |
| Other/Not Hispanic or Latinx | 0.73 | 0.35-1.53 | 0.407 |  |
| White/Hispanic or Latinx | 1.93 | 0.86-4.36 | 0.112 |  |
| Black/Hispanic or Latinx | 1.88 | 0.71-4.84 | 0.194 |  |
| Other/Hispanic or Latinx | 1.38 | 0.66-2.91 | 0.391 |  |
| Language |  |  |  |  |
| English (ref) |  |  |  |  |
| Spanish | 0.59 | 0.34-1.02 | 0.061 | 0.121 |
| Other | 0.70 | 0.34-1.4 | 0.328 |  |
| Employment |  |  |  |  |
| Employed (ref) |  |  |  |  |
| Retired | 0.53 | 0.28-0.94 | 0.037 | 0.068 |
| Unknown | 1.28 | 0.61-2.58 | 0.500 |  |
| Insurance_type |  |  |  |  |
| Private (ref) |  |  |  |  |
| Medicaid | 0.81 | 0.47-1.42 | 0.449 | 0.507 |
| Medicare | 0.69 | 0.37-1.3 | 0.249 |  |
| Pharmacy_type |  |  |  |  |
| Hospital (ref) |  |  | 0.001 | 0.001 |
| Community/retail | 0.37 | 0.26-0.52 |  |  |
| Diagnosis |  |  |  |  |
| Atrial fibrillation/flutter (ref) |  |  |  |  |
| Deep vein thrombosis | 1.05 | 0.7-1.55 | 0.824 | 0.404 |
| Pulmonary embolism | 0.61 | 0.31-1.13 | 0.125 |  |
| Other | 0.90 | 0.3-2.36 | 0.833 |  |
| Number_of_comorbidities |  |  |  |  |
| <5 (ref) |  |  |  |  |
| 5+ | 1.31 | 0.81-2.16 | 0.275 | 0.519 |
| 10+ | 1.16 | 0.67-2.04 | 0.595 |  |
| Number_of_medications |  |  |  |  |
| <5 (ref) |  |  |  |  |
| 5+ | 0.71 | 0.43-1.19 | 0.192 | 0.397 |
| 10+ | 0.70 | 0.39-1.24 | 0.217 |  |

After backwards AIC model selection (Table 4), variables associated with nonadherence were younger age (OR 1.20, CI 1.04-1.39, p=0.010), male gender (OR 1.44, CI 1.04-1.99, p=0.026), black non-Hispanic origins (OR 1.98, CI 1.17-3.47, p=0.014), employment (OR 1.92, CI 1.09-3.57, p=0.032), hospital pharmacy prescriptions (OR 2.63, CI 1.89-3.57, p=0.001), and English language (OR 1.69, CI 1.11-2.63, p=0.016). White Hispanic origin was borderline associated (OR 1.92, CI 0.91-4.04, p = 0.085). Insurance, comorbidities, and home medications were not associated.

**Table 4.**
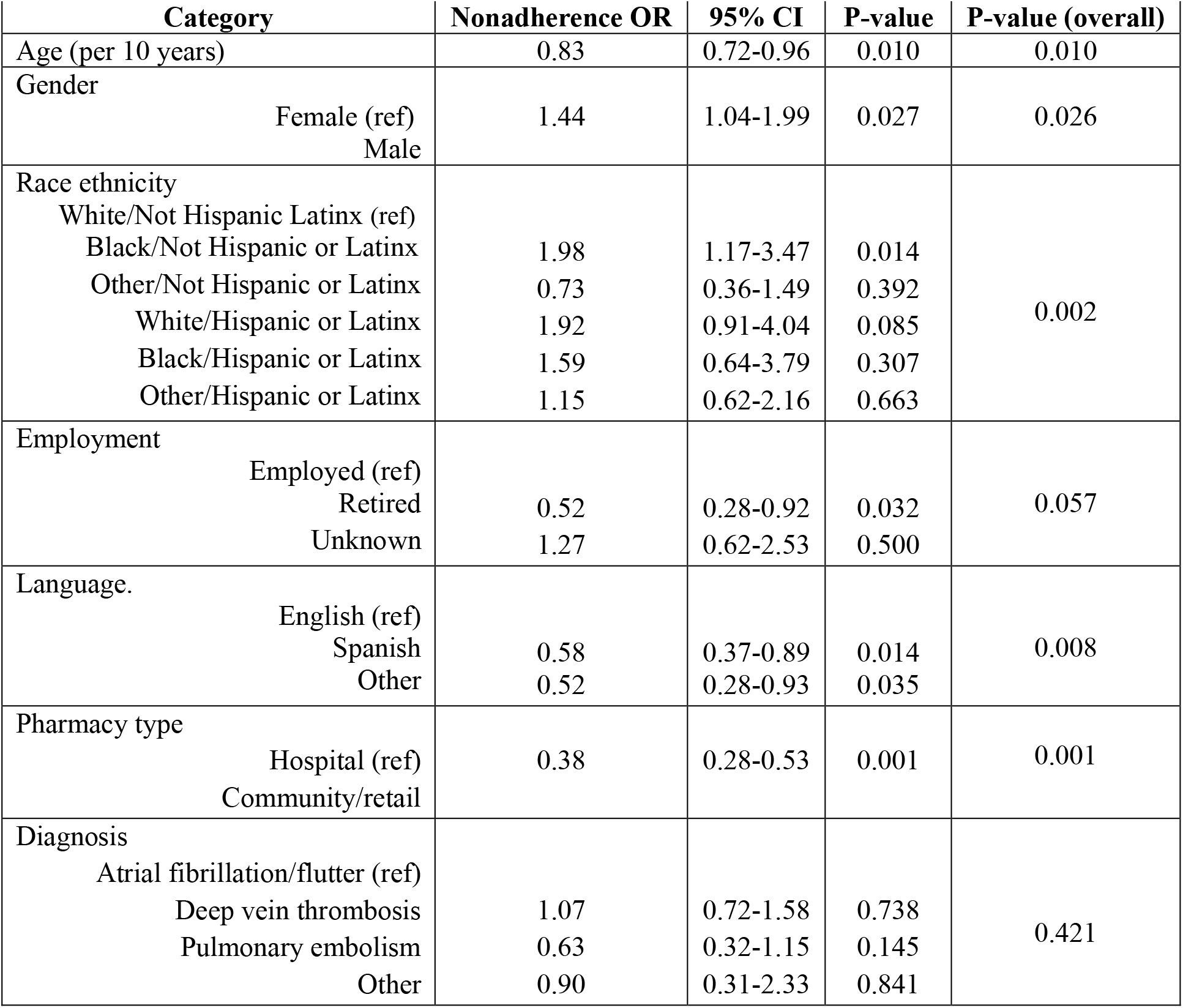
Multivariable logistic regression after backwards model selection by AIC.

| **Category** | **Nonadherence OR** | **95% CI** | **P-value** | **P-value (overall)** |
| --- | --- | --- | --- | --- |
| Age (per 10 years) | 0.83 | 0.72-0.96 | 0.010 | 0.010 |
| Gender |  |  |  |  |
| Female (ref) | 1.44 | 1.04-1.99 | 0.027 | 0.026 |
| Male |  |  |  |  |
| Race ethnicity |  |  |  |  |
| White/Not Hispanic Latinx (ref) |  |  |  |  |
| Black/Not Hispanic or Latinx | 1.98 | 1.17-3.47 | 0.014 | 0.002 |
| Other/Not Hispanic or Latinx | 0.73 | 0.36-1.49 | 0.392 |  |
| White/Hispanic or Latinx | 1.92 | 0.91-4.04 | 0.085 |  |
| Black/Hispanic or Latinx | 1.59 | 0.64-3.79 | 0.307 |  |
| Other/Hispanic or Latinx | 1.15 | 0.62-2.16 | 0.663 |  |
| Employment |  |  |  |  |
| Employed (ref) |  |  |  |  |
| Retired | 0.52 | 0.28-0.92 | 0.032 | 0.057 |
| Unknown | 1.27 | 0.62-2.53 | 0.500 |  |
| Language. |  |  |  |  |
| English (ref) |  |  |  |  |
| Spanish | 0.58 | 0.37-0.89 | 0.014 | 0.008 |
| Other | 0.52 | 0.28-0.93 | 0.035 |  |
| Pharmacy type |  |  |  |  |
| Hospital (ref) | 0.38 | 0.28-0.53 | 0.001 | 0.001 |
| Community/retail |  |  |  |  |
| Diagnosis |  |  |  |  |
| Atrial fibrillation/flutter (ref) |  |  |  |  |
| Deep vein thrombosis | 1.07 | 0.72-1.58 | 0.738 | 0.421 |
| Pulmonary embolism | 0.63 | 0.32-1.15 | 0.145 |  |
| Other | 0.90 | 0.31-2.33 | 0.841 |  |

Spanish speakers had lower odds of nonadherence than English speakers while Hispanics had higher odds of nonadherence compared to non-Hispanic Whites. To delineate the effect of Spanish speakers vs English speakers from race/ethnicity, we performed an analysis for nonadherence using the combined race/ethnicity and language variable (e.g., Hispanic White who speaks Spanish vs Hispanic White who speaks English). In Table 5, compared to non-Hispanic Whites who speak English, English-speaking non-Hispanic Blacks, Hispanic/Whites, and Hispanic/Blacks showed increased odds of nonadherence (OR=1.23, 1.40, 1.18, respectively), while English-speaking non-Hispanic/other had lower odds of nonadherence (OR=0.48). Compared to English speaking non-Hispanic Whites, Spanish speaking Hispanic Whites and Blacks showed no notable difference in nonadherence (OR=0.99 and 0.99), while non-Hispanic/other individuals had lower odds of nonadherence (OR=0.2) than Hispanic/other individuals (OR=0.62). We observed that Spanish speakers showed lower odds of nonadherence than English speaking individuals (Table 6) consistently across race/ethnicity groups (OR=0.41, 0.71, 0.84, 0.61 among non-Hispanic/other, Hispanic/white, Hispanic/black, Hispanic/other respectively).

**Table 5.**
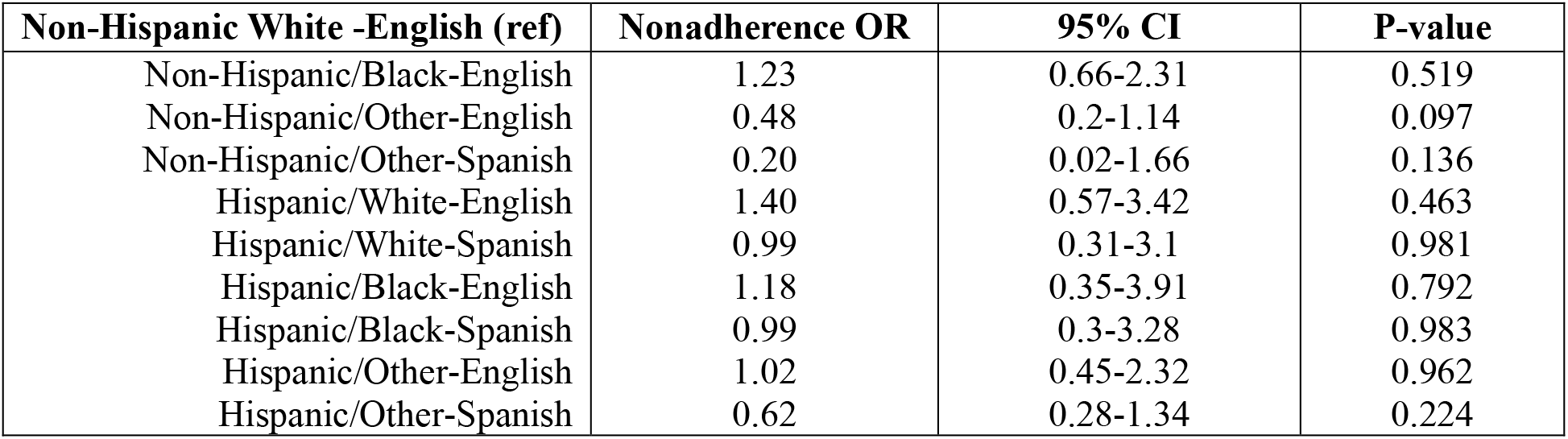
Association between combined race/ethnicity and primary language with nonadherence, adjusted for age, gender, employment, and pharmacy using OR and CI.

| **Non-Hispanic White -English (ref)** | **Nonadherence OR** | **95% CI** | **P-value** |
| --- | --- | --- | --- |
| Non-Hispanic/Black-English | 1.23 | 0.66-2.31 | 0.519 |
| Non-Hispanic/Other-English | 0.48 | 0.2-1.14 | 0.097 |
| Non-Hispanic/Other-Spanish | 0.20 | 0.02-1.66 | 0.136 |
| Hispanic/White-English | 1.40 | 0.57-3.42 | 0.463 |
| Hispanic/White-Spanish | 0.99 | 0.31-3.1 | 0.981 |
| Hispanic/Black-English | 1.18 | 0.35-3.91 | 0.792 |
| Hispanic/Black-Spanish | 0.99 | 0.3-3.28 | 0.983 |
| Hispanic/Other-English | 1.02 | 0.45-2.32 | 0.962 |
| Hispanic/Other-Spanish | 0.62 | 0.28-1.34 | 0.224 |

**Table 6.** Association between primary language and race/ethnicity with nonadherence using OR and CI.

| **Spanish vs English** | **Nonadherence OR** | **95% CI** | **P-value** |
| --- | --- | --- | --- |
| Among Non-Hispanic/Other | 0.41 | 0.05-3.53 | 0.418 |
| Among Hispanic/White | 0.71 | 0.21-2.35 | 0.569 |
| Among Hispanic/Black | 0.84 | 0.19-3.71 | 0.818 |
| Among Hispanic/Other | 0.61 | 0.28-1.31 | 0.201 |

## Discussion

Former studies on AC therapy have identified multiple clinical factors that influence adherence including but not limited to adverse effects, restrictions on food and medication intake, requirement for routine monitoring, and dosing frequency^1,2^. Although the implementation of DOACs has largely eliminated clinical barriers, patient and systemic level demographic factors continue to hinder adherence. Among these factors are the time constraints of intake, complexities in medication dosing, medication expenses, lack of insurance coverage, fear of no antidote, and workplace financial repercussions^2^. Other barriers in adherence include cognitive impairment, dementia, poor knowledge, lack of patient provider goal agreement, medication shortages in pharmacies, and conflicting recommendations from different providers^6^. Overall, factors have been grouped as therapy-related, patient-related, healthcare provider-related, and health system-related^6^.

This retrospective review of DOAC intake in the Bronx found significant correlations between nonadherence and factors such as young age, male gender, black non-Hispanic origins, hospital pharmacy based prescriptions, employed status, and English as primary language. Several reasons for these associations can be inferred, although they would warrant further investigation with an individual level study. The association between nonadherence and younger age could on one hand be attributed to younger patients being generally healthier than older patients and therefore less likely to experience significant symptoms or mortality when deviating from recommended treatments. On the other hand, younger patients may perceive themselves to be healthy and thereby deviate from provider guidelines. Another possibility is that younger patients may be less aware of the implications of their diagnosis and the burden of health problems whereas older patients who have experienced health problems and symptoms for a long time are more aware and understand the importance of adherence to treatment.

The association between nonadherence and male gender could have several explanations which too would require individual level analysis. One explanation could be that males perceive themselves to be healthier than do females and are thus less likely to follow established guidelines. Alternatively, males may be less debilitated by symptoms or to seek health services if ill and thereby less likely to utilize pharmacotherapy in a routine pattern. In both scenarios, males are more likely to exercise independent decisions regarding treatment than are females and therefore deviate from recommended treatment regimens. Finally, males may have a lesser understanding of their therapy as compared to females and therefore be more nonadherent.

The correlation between nonadherence and hospital pharmacy based prescriptions was quite significant. One explanation could be the small ratio of hospital pharmacies to patient population. Notably, there are less than 10 hospital based pharmacies serving close to 1000 DOAC patients in the Bronx whereas retail pharmacies number at least 50-60 in the same area. Moreover, retail pharmacies may be more likely to face competition from co-existing retail pharmacies and therefore be more likely to ensure medication availability. Finally, the length of time over which services are available per day could also explain the gap in adherence; many retail pharmacies are open nearly 24 hours a day while hospital pharmacies have limited hours in the day and such flexibility works to the advantage of the former.

The association between nonadherence and being employed could be explained on the basis that employment provides patients less time to focus on their health and treatment. Patients who are retired are free of work related stresses and time constraints, and thus can focus more on their health.

Generally, English speaking capacity is expected to translate into improved communications and increased ability of patients to understand provider guidance both of which are difficult to achieve with non-English speakers. Ironically, nonadherence was more frequently observed among English than Spanish speakers in our study. This paradoxical finding suggests a few possibilities. One could be that English speakers are more familiar with health systems and confident in making health decisions independent of provider guidance while Spanish speakers may be less familiar with systems and more dependent on provider guidance. This discrepancy in adherence warrants further study into whether English is utilized by native populations while foreign languages are utilized by immigrant populations and if natives may be more nonadherent than immigrants. Alternatively, the discrepancy between English and Spanish speakers may have to do with the fact that English speakers comprised a large proportion of black non-Hispanics. Nonadherence was notably more common in this racial ethnic population than in others, potentially linking English speaking ability with higher rates of nonadherence.

## Limitations

One of the limitations in our study was the size of the patient population. Considering that the patient population in the Bronx numbers more than 10,000 but the cohort was only about 1000 patients, this population size limited the power of our study and ability to determine how insurance type, diagnosis type, comorbidities, home medications, subgroups in race, and other categories contribute to nonadherence. Moreover, among the categories that were significant, most correlations were weakly significant indicating the need for a larger study base. A second limitation was the lack of data on education status. Level of education and knowledge about medications are known to play roles in adherence but without this information, further stratification of the data and correlations based on education level was not possible. A third limitation was the absence of individual level reasons for nonadherence. The data for the study utilized demographic characteristics listed in the electronic records to make correlations to nonadherence. Patients’ individual perspectives on barriers to adherence were not acquirable. Further studies with responses from individuals about DOAC habits would delineate the underlying reasons for the associations in our study and challenges that need to be addressed to improve adherence.

## Conclusion

DOAC nonadherence in the Bronx has significant correlations with young age, male gender, black non-Hispanic origins, hospital pharmacy based prescriptions, active employment, and English as a primary language. Nonadherence did not significantly correlate with patient insurance type, diagnosis type, comorbidity burden, or number of home medications. However due to borderline significance seen with some of the non-correlating categories, future investigations utilizing a larger population base may elucidate the true connections between these categories and DOAC nonadherence.

## Data Availability

All data are available in the article and online supplement

## Abbreviations

DOAC: direct oral anticoagulation
INR: international normalized ratio
HIT: heparin induced thrombocytopenia
DVT: deep vein thrombosis
PE: pulmonary embolism
OR: odds ratio
CI: confidence interval
AF: atrial fibrillation/flutter
AC: anticoagulation
AIC: Akaike Info Criterion

